# A blinded, counterbalanced rater design for evaluating AI-assisted summarisation of tertiary clinical genomics reports: methodology of the QNOMX-VHIR-CPSP-001 Phase 1 study

**DOI:** 10.64898/2026.06.11.26355467

**Authors:** James Creeden, Marcus Olivecrona, Aroa Soriano

**Author notes:** **Corresponding author:** James Creeden MD PhD, Qnomx AG, Switzerland Innovation Park Basel Area AG, Lichtstrasse 35, 4056 Basel, Switzerland.

## Abstract

**Background:** Tertiary clinical genomics reports condense layered molecular findings into documents that treating oncologists must read, translate, and act upon; manual summarisation of these reports is time-consuming and variable. Tools that assist summarisation and translation into local languages are emerging, yet the field lacks an agreed methodology for evaluating such tools before any downstream clinical use. The appropriate first endpoint is fidelity of the generated summary to its source report, assessed by qualified human raters under blinded scoring, not downstream variant classification.

**Methods:** QNOMX-VHIR-CPSP-001 Phase 1 is a single-site, non-interventional clinical performance study conducted at Vall d’Hebron Institut de Recerca (VHIR) under ISO 20916:2019 as a Clinical Performance Study Protocol. De-identified tertiary cancer genomics reports from pediatric oncology cases are summarised by the AI-assisted summarisation system under evaluation and, in parallel, by the standard manual workflow. Qualified raters score both summary types against the source genomics report using the Quality Summary Index (QSI), a six-dimension, five-point rubric adapted from the Provider Documentation Summarization Quality Instrument, under a blinded, counterbalanced, two-period crossover with a minimum fourteen-day washout. Two co-primary composite endpoints, content and presentation, are analysed for non-inferiority under a Bayesian hierarchical model, with a frequentist linear mixed model as the convergence check. Inter-rater reliability is reported as Krippendorff’s α; a Monte-Carlo power analysis of the fixed clustered design is pre-specified.

**Discussion:** The design isolates summarisation quality from clinical decision-making by scoring both summary types against the same source report under blinding, counterbalancing, and a fourteen-day washout.

**Conclusion:** The QSI rubric, the counterbalanced crossover, and the pre-specified Bayesian primary with frequentist convergence check define a replicable protocol for early-stage evaluation of AI-assisted summarisation in tertiary genomics reporting; observed variance components will inform sample-size determination for Phase 2.

## 1. Introduction

Precision oncology now depends on tertiary genomics reports: integrated molecular documents that consolidate sequencing results, copy-number findings, fusion calls, and the variant classifications and therapeutic annotations attached to them. These reports are dense, frequently issued in English, and written for specialist molecular tumour boards; the community oncologist who must use these laboratory results at the point of care needs a concise, accurate summary, and today that summary is produced in laboratories by hand, often translated into local language for the treating physician. The task is time-consuming, and outputs are highly variable in completeness and clarity. A community oncologist parsing a 10–20 page, dense genomics report [2] between patient encounters is frequently working without subspecialty training in precision oncology with rapidly evolving guidelines [4]. Distinguishing a clinically actionable alteration from variants of uncertain significance, identifying a germline alteration that indicates a genetic counseling referral, and prioritising among listed treatment options all depend on expertise that the report’s format assumes but the clinical workflow does not guarantee [5].

Systems that assist this summarisation are now technically feasible, and the literature on machine assistance in clinical genomics has matured around the adjacent problems of evidence retrieval and variant curation pipeline support [6,7,8]. Yet the evaluation methodology has not kept pace with the tools themselves. Two studies of human evaluation of large language models in healthcare document recurring deficiencies, including absent sample-size justification, under-specified rater numbers and training, and metrics chosen without a psychometric basis [9,10].

The failure modes that matter when a tertiary genomics report is condensed are specific: the omission of a clinically relevant finding, the introduction of an unsupported assertion, the loss of relevance, and the degradation of language for the intended reader. These are properties of the relationship between a summary and its source, and they map onto the faithfulness problem that the clinical natural-language-generation community has begun to formalise for long-form medical summarisation [11,12].

Content fidelity is the degree to which a summary represents its source without addition, distortion, or material omission. It is observable, scorable against a fixed reference, and the property a clinician reader most needs to trust. An endpoint at this stage must isolate the summarisation step itself: the variant classifications already exist in the source tertiary report, so a classification-accuracy endpoint would measure the upstream laboratory pipeline rather than the summarisation step; and anchoring early evaluation to clinical-decision outcomes would assume the clinical-use context that this non-interventional design excludes.The QSI instrument draws on two established traditions. The ACMG/AMP 2015 standards establish that interpretive statements about genomic findings must be traceable to defined evidence categories and applied reproducibly [13]; ClinGen has operationalised that principle through curation standard operating procedures and structured expert review [14]. The Physician Documentation Quality Instrument (PDQI-9) established that note quality can be decomposed into anchored, rated attributes with measurable inter-rater reliability [15]; the Provider Documentation Summarization Quality Instrument (PDSQI-9) extended that framework to the evaluation of large-language-model-generated clinical summaries and validated it on real-world multi-document data [16].

The QSI adapts the PDSQI lineage to the genomics summarisation task: it decomposes quality into defined dimensions with behavioural anchors, scored by trained raters, so that agreement can be measured and the instrument can be applied at other sites and in other healthcare contexts.

## 2. Methods

### 2.1 Study design

QNOMX-VHIR-CPSP-001 is a non-interventional clinical performance study, documented as a Clinical Performance Study Protocol (CPSP) under EN ISO 20916:2019 [1]. The standard was chosen as the methodological reference for protocol structure, bias control through blinding and counterbalancing, monitoring, traceability, and reporting; its use governs study conduct and does not assert a regulatory classification of the system under evaluation. The quality of generated summaries relative to their source reports was assessed using the QSI instrument described in Table 1, adapted from the PDSQI-9 framework [16]. The present report describes a research evaluation in which outputs of the system under evaluation were not used for patient management; the manual standard-of-care summary remains the only clinical artefact. Phase 1 is a retrospective evaluation: de-identified archived reports are summarised, the summaries are scored under blinding, and the resulting QSI scores characterise summarisation quality. The design is a blinded, counterbalanced, two-period crossover at the level of the rater-case pair.

**Table 1.** QSI dimensions, definitions, and scoring anchors.

| # | Dimension | What it assesses (against the source report) | Anchor logic (5 to 1) |
| --- | --- | --- | --- |
| 1 | Accuracy | Factual correctness of stated information relative to the source | 5 = no errors; 4 = one minor inaccuracy; 3 = multiple minor; 2 = one major; 1 = multiple major errors |
| 2 | Completeness | Inclusion of all clinically relevant findings present in the source | 5 = all findings; 4 = one minor omission; 3 = multiple minor; 2 = one major; 1 = multiple major omissions |
| 3 | Relevance | Focus on actionable and key findings for the intended user | 5 = no irrelevancies; graded to 1 = multiple major irrelevancies |
| 4 | Organisation | Logical grouping and ordering of information | 5 = appropriate throughout; graded to 1 = inappropriate |
| 5 | Language | Clarity, correctness, and professional register for the intended reader | 5 = clear and correct; graded to 1 = pervasive errors and unclear |
| 6 | Succinctness | Brevity without loss of key information or redundancy | 5 = minimal, non-redundant; graded to 1 = excessive wordiness |

For each case, both a manual summary and a summary produced by the system under evaluation are scored by the same raters; raters are blinded to the origin of each summary, and the order in which a rater encounters the two summary types for a given case is counterbalanced across cases, with a washout interval between the two periods to attenuate recall and halo effects (ISO 20916:2019, B.8.1 c)) [1].

### 2.2 Setting and oversight

The study is conducted at VHIR, Barcelona, with Qnomx AG, Basel, as sponsor. Source material derives from archived de-identified reports generated during routine clinical care within the COMIK (Instituto de Salud Carlos III: PI21/01661) and SEHOP-PENCIL (Instituto de Salud Carlos III: PMP21/00073) research projects. Governance follows the responsibilities defined in the CPSP, including sponsor accountability for data quality and integrity, investigator responsibility for source documentation and access for monitoring, and risk-based monitoring.

Ethics oversight is provided by the Comitè d’Ètica en la Investigació amb Medicaments (CEIM) at VHIR. A favourable CEIM opinion was obtained under reference PR(AMI)318-2025 (Dra. Soriano), dated 08 August 2025, including a waiver of additional informed consent for the use of de-identified data from the COMIK and SEHOP-PENCIL projects in this non-interventional design (ISO 20916:2019, 4.1 to 4.5) [1].

### 2.3 Materials: source reports, summaries, and blinding

Study inputs are tertiary cancer genomics reports and the related case documentation required to produce a standard manual summary, derived from pediatric oncology cases (ISO 20916:2019, 5.5.3.9) [1]. Pediatric oncology was selected for VHIR institutional expertise, the availability of curated retrospective datasets, and the clinical need for efficient genomics summarisation in pediatric precision oncology. Cases are eligible where the source report and supporting documentation permit adequate scoring under the QSI; cases are excluded where documentation is insufficient for fair comparison, or where de-identification deficits cannot be remediated before processing.

For each eligible case, a VHIR coordinator de-identifies the source report under a documented redaction procedure and assigns an anonymised case identifier; the system under evaluation then generates a summary within a controlled environment that records audit elements including software, model, and prompt versions, timestamps, and user role. Blinding is implemented at the artefact level: each summary presented for scoring is relabelled with a round-specific pseudonym so that raters cannot infer the summary’s origin and cannot link the two summary types for the same case across periods.

### 2.4 Raters: eligibility, training, and calibration

Raters are qualified oncologists with at least four years of experience in cancer genomics interpretation. The Phase 1 rater roster comprises three raters: one rater scores every report, and two further raters each score a complementary, non-overlapping subset, so that each of the 38 analysis cases receives two independent ratings (76 paired observations). Before study scoring, all raters complete a documented training workshop covering the QSI rubric, its dimensions, and scoring guidance, followed by independent scoring of a familiarisation set of eight training cases (presented in the system-output arm, several with deliberately introduced errors to exercise rater vigilance for omission and unsupported content), used to standardise application of the rubric; these eight cases are excluded from all inference, consistent with psychometric practice for familiarisation scoring sets [15,16]. Inter-rater reliability for the study scoring is reported as Krippendorff’s α as the primary metric, with pairwise quadratic-weighted Cohen’s κ, Spearman ρ, and exact-agreement percentage for overlapping rater pairs, reflecting the unbalanced rater roster (Statistical Analysis Plan, Section 5.3).

### 2.5 The QSI rubric and the content-fidelity quality attribute

The Quality Summary Index (QSI) is a structured rater instrument that assesses a genomics summary against its source report across six dimensions, each scored on a five-point Likert scale anchored from 1 (unacceptable) to 5 (excellent). The instrument is adapted from the PDSQI-9 and its PDQI-9 antecedent [15,16], with dimension definitions and anchors specialised for tertiary genomics reporting. The dimensions and their anchoring logic are summarised in Table 1.

Accuracy and Completeness govern what the summary asserts relative to the source: whether each stated fact is correct, and whether every clinically relevant finding survives the summarization. Relevance, Organisation, Language, and Succinctness govern how the retained content is presented: whether the summary includes clinically relevant findings, groups them coherently, expresses them clearly for the intended reader, and does so without redundancy.

Content fidelity is operationalised principally through Accuracy and Completeness, with Relevance bearing on the judgement of what counts as material. Quality is summarised by two composite scores defined as the means of the underlying dimensions on the 1–5 scale: a content composite (accuracy, completeness, relevance) and a presentation composite (organisation, language, succinctness); an overall composite is the mean of all six dimensions.

Succinctness is not a stylistic preference and warrants particular emphasis in the genomics setting. The reader of a tertiary genomics summary is typically a community oncologist working under acute time constraints, for whom attention is a very limited resource; a summary that buries the two or three decision-relevant findings inside paragraphs of restated detail imposes the same cognitive burden that the summarisation was meant to relieve. The documentation-quality literature treats brevity as a measurable attribute for this reason, and Croxford et al. found that input note length correlated negatively with Succinctness (ρ = −0.200, p = 0.029) and Organisation (ρ = −0.190, p = 0.037) scores [16], consistent with the observation that length and signal are in tension once the essential content is present. In genomics specifically, where a single actionable alteration or a germline-referral trigger can be decisive, succinctness determines whether content fidelity actually reaches the reader’s attention rather than merely residing somewhere in the text.

### 2.6 Counterbalancing schema and randomisation

The Phase 1 design separates a small set of calibration and training cases from the cases that enter the randomised crossover. The remaining 38 cases enter a two-period crossover in which each case is scored by two of three raters, so that every case receives exactly two independent evaluations and each rater scores both summary types for each assigned case across the two periods.

Case-to-rater assignment is deterministic. One rater scores all 38 analysis cases, and the other two raters each score one of two complementary 19-case subsets, with the subset assignment generated by a single deterministic shuffle of the 38 cases. Modality order is counterbalanced through a case-level batch property: the 38 analysis cases are split into two batches of 19, with batch 1 cases scored in the system-output arm in round 1 and the manual arm in round 2 and batch 2 cases scored in the reverse order; the case-to-batch assignment is generated by the same deterministic shuffle. This structure counterbalances arm against round at the aggregate level, so that any round, learning, or fatigue effect is orthogonal to the arm contrast. The implemented tables achieve a global balance of 38 manual-first and 38 system-first orders across the 76 rater-case pairs and are documented and signed before study scoring.

Randomisation was performed with the Python standard-library random.Random.shuffle; the seed, tool, and date are recorded in the controlled randomisation log.

A minimum fourteen-day calendar washout is enforced between a rater’s two scoring events for the same case, to allow decay of recall and to attenuate halo effects (ISO 20916:2019, B.8.1 c)) [1]; washout completion dates are tracked in the master randomisation table, and a shorter interval is logged as a protocol deviation. The two scoring events for the same rater-case pair fall in different rounds; the fourteen-day calendar interval is enforced in addition to the round separation. Round-specific pseudonyms prevent a rater from linking the two summary types for a case across periods. Scoring is captured on timestamped electronic forms with an immutable audit log.

### 2.7 Statistical analysis plan

The analysis sets follow the statistical analysis plan (SAP). The per-protocol set comprises cases meeting eligibility, successfully processed, and with complete scoring of both summary types by the required number of raters and without major deviation; the intention-to-treat set comprises all enrolled cases with at least one QSI score.

The primary analysis is a non-inferiority comparison of the system summary against the manual summary on two co-primary composite endpoints on the 1–5 QSI scale: a content composite (mean of accuracy, completeness, relevance) and a presentation composite (mean of organisation, language, succinctness). For each, the non-inferiority margin is m = 0.5 composite points, tested one-sided at α = 0.025; non-inferiority is concluded where the lower bound of the two-sided 95% interval for Δ = mean(system − manual) lies above −m. Both co-primary endpoints must succeed (intersection–union); no multiplicity adjustment is applied across the two co-primary tests. The non-inferiority margin of 0.5 composite points is a pragmatic threshold anchored to the structure of the QSI scale: the composite is the mean of three dimensions scored on a 1–5 grade interval, and a composite-mean decrement of 0.5 corresponds on average to a half-grade drop on each of the three dimensions in the composite. We do not claim that 0.5 is the smallest difference worth excluding in the sense of EMA CHMP/EWP/2158/99 [17]; we therefore report the posterior of Δ together with non-inferiority decisions at both the 0.5 margin and a stricter 0.25-point benchmark, and interpret accordingly.

The primary estimator is a Bayesian hierarchical model with a Gaussian likelihood on the composite, fixed effects for arm and round, and crossed random effects for case, rater, and case-by-arm. The fixed effects (intercept, arm, round) are given Normal(0, 1) priors and all standard-deviation parameters - the residual SD and the case, rater, and case-by-arm SDs - HalfNormal(1) priors. These are weakly informative on the 1–5 composite scale: a Normal(0, 1) prior on the arm effect is centred at zero (no *a priori* preference for either arm, as appropriate under a non-inferiority framing) yet places substantial mass well beyond any clinically plausible shift - its scale of one full composite point is twice the 0.5-point margin - so it constrains the geometry without driving the estimate; the HalfNormal(1) priors concentrate the variance components at the modest dispersion expected when scores cluster high while keeping them strictly positive, regularising the weakly identified rater SD (three levels) and case-by-arm SD (38 levels) away from the zero boundary at which an unpenalised maximum-likelihood fit can collapse. The three random effects use a non-centered parameterisation (u = σ · u_raw, u_raw ∼ Normal(0, 1)), which is mathematically equivalent to the centered form but yields the well-conditioned geometry that, together with a high NUTS target acceptance probability of 0.99, eliminates the funnel divergences that otherwise arise under wide priors. The model is fitted via the No-U-Turn sampler [18] with four chains of 1000 warmup and 1000 retained draws each (4000 posterior samples); convergence is required at 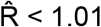 with zero divergences. The non-inferiority decision is the resulting posterior statement (the 95% credible interval for Δ and P(Δ > −m)) and the credible interval integrates the uncertainty in the case-by-arm variance into the interval for Δ, whereas a Wald interval treats that estimated variance as known. As the pre-specified sensitivity analysis, a frequentist linear mixed model is fitted to the within-(case × rater) paired differences, in which the case and rater main effects cancel by construction; because the design delivers complete, balanced pairs this model is expected to closely reproduce the primary estimate, and the primary conclusion stands only where the two agree in direction. Robustness to the prior is verified by a pre-specified widening of the priors, in which the fixed-effect SD and all HalfNormal scales are inflated by a common factor (1.0, 2.5, 5.0); a non-inferiority verdict stable in Δ^^^ and P(Δ > −m) across these scales demonstrates the conclusion is not an artefact of the prior.

Secondary analyses comprise non-inferiority on each of the six QSI dimensions with Holm adjustment, two-sided superiority tests on each composite as exploratory, the round fixed effect as a check on the crossover counterbalancing, and per-rater arm means as a check that the contrast is not driven by a single rater. Inter-rater reliability is reported as Krippendorff’s α with an ordinal difference function for the raw 1–5 dimensions and an interval function for the continuous composites [19]. Pairwise inter-rater agreement is reported as quadratic-weighted Cohen’s κ, Spearman ρ, and exact-agreement percent.

### 2.8 Sample size and power

The sample size was determined by the curated material available: 38 eligible analysis cases were included, yielding 76 paired observations per endpoint. A pre-specified Monte-Carlo power analysis of the fixed clustered design was conducted to confirm that this feasibility-determined sample is not materially underpowered for the primary comparison. Simulating the actual design with a case-cluster-robust standard error under truly equivalent arms, the study has ≥ 0.99 power to declare non-inferiority at the m = 0.5 margin (one-sided α = 0.025) on each co-primary composite across the plausible parameter range (paired-difference SD ≤ 0.8; intra-case correlation ≤ 0.5); power at the stricter 0.25-point margin is adequate only when the paired-difference SD is ≤ 0.6. Because this is a design-stage check, the power analysis is not re-computed on the observed data; the observed variance components will instead inform the sample-size justification for the planned Phase 2.

### 2.9 Data management, monitoring, and deviation handling

Data management follows a documented data management plan specifying query handling, validation of electronic systems, access controls, audit trails without deletion of entered data, backup, and a ten-year retention period.

Risk-based monitoring verifies study conformity, training completion, data completeness, deviation handling, and traceability of both data and software configuration, with the extent of source data verification and the monitoring modality set out in the monitoring plan.

Deviations are recorded, graded, and escalated under a deviation and corrective-and- preventive-action procedure; for this software study the device-deficiency definitions, specified in the CPSP as study-specific applications of the ISO framework, explicitly include critical omission of findings, unsupported content presented as fact, corrupted export, and audit-log or access-control failure (ISO 20916:2019, 3.16, 5.5.3.19, and B.8.11) [1].

## 3. Discussion

This design isolates the summarisation quality from clinical decision-making by scoring both summary types against the same source report under blinding, counterbalancing, and washout. It measures content fidelity directly and removes several well-described sources of rater bias. The inclusion of training cases with deliberately introduced errors exercises rater vigilance for omission and unsupported content, the failure modes that carry the highest stakes in genomics summarisation. The QSI is a six-dimension, anchored rubric, with rater agreement reported as Krippendorff’s α together with pairwise quadratic-weighted Cohen’s κ, Spearman ρ, and exact-agreement percentage. It is applied here to pediatric oncology at a single site and structured to be adaptable to other genomics reporting contexts; transferability is the subject of planned multi-site work.

The design is a protocol-stage, non-interventional study under ISO 20916; it pre-specifies the endpoints, the analysis, and the bias controls without committing the system under evaluation to any clinical use. Reporting of the observational structure follows STROBE [21]; DECIDE-AI [20] addresses the subsequent prospective decision-support stage, which is the scope of Phase 2. The choice of expert human assessment as the reference standard for judging generated clinical text follows the convention established in clinical-summarisation evaluation [12,22].

Several limitations follow from the scope. The study is single-site with one curated dataset; rubric behaviour and the agreement it yields may not transfer unchanged to other centres, to adult oncology, or to non-oncology genomics. It evaluates one system, so the design characterises an instrument applied to a single tool, not a comparison across tools. The sample size was determined by the curated material available rather than by a power calculation; a non-significant result should be read as absence of demonstrated non-inferiority, not evidence of equivalence.

The QSI is adapted from PDSQI-9 [16] for the genomics setting; rater training standardises its application, but content validity for the specific failure modes of tertiary genomics summarisation has not been independently established and is an objective for subsequent validation work.

Inter-rater reliability is constrained by the three-rater design with one rater scoring all 38 cases; Krippendorff’s α is therefore conditional on that rater’s variance and cannot fully separate idiosyncratic scoring from rater-pair disagreement. We treat the composite as continuous under a Gaussian likelihood; this is a tractability choice and a cumulative-link analysis at the dimension level may strengthen subsequent reports. Blinding integrity is supported by round-specific pseudonyms and a controlled scoring environment but is not formally assessed; given the human-versus-machine contrast under test, raters may retain residual ability to infer summary origin from stylistic features. The non-inferiority margin of 0.5 composite points is pragmatic rather than empirically validated; conclusions should be read against the 0.25-point benchmark. Content fidelity is a necessary but not sufficient condition for clinical utility: the present design is deliberately silent on operational use and on downstream classification and decision outcomes, which lie outside the scope of a non-interventional evaluation.

Evidence retrieval and curation support [6,7,8] and variant-interpretation standards [13,14] address steps upstream of a tertiary genomics report. The QSI addresses a different and less-studied step: whether a condensed representation of an already-issued report preserves the information a clinician reader needs, and preserves it succinctly enough to be used.

## Data Availability

The QSI rubric and the randomisation and counterbalancing procedure are available from the corresponding author on reasonable request.

## 4. Declarations

### 4.1 Ethics approval and consent

The study is governed by the CEIM at VHIR; a favourable opinion including a waiver of additional informed consent for the non-interventional use of de-identified archived data was obtained under reference PR(AMI)318-2025, dated 08 August 2025. The study is conducted in accordance with the Declaration of Helsinki [23] and applicable data-protection law, including the EU GDPR [24] and the Swiss FADP; the de-identification procedure, the Data Protection Impact Assessment, and the legal basis for data transfer are specified in the study data-protection documentation.

### 4.2 Funding

The study is sponsored by Qnomx AG, Basel, Switzerland. Source material derives from the COMIK and SEHOP-PENCIL research projects at VHIR, funded by the Instituto de Salud Carlos III through grants PI21/01661 and PMP21/00073 (co-funded by the European Regional Development Fund/European Social Fund; “A way to make Europe”/”Investing in your future”).

### 4.3 Competing interests

J.C. and M.O. are each co-founders and employees of Qnomx AG and hold equity in Qnomx AG; Qnomx AG is the sponsor of this study, and the AI-assisted summarisation system under evaluation is a Qnomx product. A.S. declares no competing interest.

### 4.4 Data availability

The QSI rubric and the randomisation and counterbalancing procedure are available from the corresponding author on reasonable request. De-identified source reports and individual rater scores are governed by the CEIM approval and applicable data-protection law and are held under controlled access at VHIR with a ten-year retention period; they are not publicly available.

### 4.5 Author contributions (CRediT)

J.C.: Conceptualization; Methodology; Investigation; Writing, original draft. M.O.: Data curation; Formal analysis; Software; Validation; Methodology. A.S.: Conceptualization; Methodology; Resources; Supervision; Project administration; Writing, review and editing. All authors approved the final manuscript.

## Acknowledgements

We thank Asbleidy Carolina Torres for project coordination, regulatory management and support with ethics approval and data protection procedures. We thank the raters for their contribution to the scoring process.

## Notes

### Author Declarations

Ethics committee Comitè d’Ètica en la Investigació amb Medicaments (CEIM) of Vall d'Hebron Research Institute (VHIR), Universitat Autònoma de Barcelona, Barcelona, Spain gave ethical approval for this work under reference PR(AMI)318-2025 (Dra.Soriano), dated 08 August 2025, including a waiver of additional informed consent for the use of de-identified data from the COMIK and SEHOP-PENCIL projects in this non-interventional design.

